# Automated detection and staging of malaria parasites from cytological smears using convolutional neural networks

**DOI:** 10.1101/2021.01.26.21250284

**Authors:** Mira S. Davidson, Sabrina Yahiya, Jill Chmielewski, Aidan J. O’Donnell, Pratima Gurung, Myriam Jeninga, Parichat Prommana, Dean Andrew, Michaela Petter, Chairat Uthaipibull, Michelle Boyle, George W. Ashdown, Jeffrey D. Dvorin, Sarah E. Reece, Danny W. Wilson, D. Michael Ando, Michelle Dimon, Jake Baum

## Abstract

Microscopic examination of blood smears remains the gold standard for diagnosis and laboratory studies with malaria. Inspection of smears is, however, a tedious manual process dependent on trained microscopists with results varying in accuracy between individuals, given the heterogeneity of parasite cell form and disagreement on nomenclature. To address this, we sought to develop an automated image analysis method that improves accuracy and standardisation of cytological smear inspection but retains the capacity for expert confirmation and archiving of images. Here we present a machine-learning method that achieves red blood cell (RBC) detection, differentiation between infected and uninfected RBCs and parasite life stage categorisation from raw, unprocessed heterogeneous images of thin blood films. The method uses a pre-trained Faster Region-Based Convolutional Neural Networks (R-CNN) model for RBC detection that performs accurately, with an average precision of 0.99 at an intersection-over-union threshold of 0.5. A residual neural network (ResNet)-50 model applied to detect infection in segmented RBCs also performs accurately, with an area under the receiver operating characteristic curve of 0.98. Lastly, using a regression model our method successfully recapitulates intra-erythrocytic developmental cycle (IDC) stages with accurate categorisation (ring, trophozoite, schizont), as well as differentiating asexual stages from gametocytes. To accelerate our method’s utility, we have developed a mobile-friendly web-based interface, PlasmoCount, which is capable of automated detection and staging of malaria parasites from uploaded heterogeneous input images of Giemsa-stained thin blood smears. Results gained using either laboratory or phone-based images permit rapid navigation through and review of results for quality assurance. By standardising the assessment of parasite development from microscopic blood smears, PlasmoCount markedly improves user consistency and reproducibility and thereby presents a realistic route to automating the gold standard of field-based malaria diagnosis.

**Significance Statement:** Microscopy inspection of Giemsa-stained thin blood smears on glass slides has been used in the diagnosis of malaria and monitoring of malaria cultures in laboratory settings for >100 years. Manual evaluation is, however, time-consuming, error-prone and subjective with no currently available tool that permits reliable automated counting and archiving of Giemsa-stained images. Here, we present a machine learning method for automated detection and staging of parasite infected red cells from heterogeneous smears. Our method calculates parasitaemia and frequency data on the malaria parasite intraerythrocytic development cycle directly from raw images, standardizing smear assessment and providing reproducible and archivable results. Developed into a web tool, PlasmoCount, this method provides improved standardisation of smear inspection for malaria research and potentially field diagnosis.

## Introduction

Malaria is an infectious disease caused by protozoan parasites from the genus *Plasmodium*, of which *Plasmodium falciparum* is the most common and lethal to humans (1). Despite advances in rapid point-of-care diagnostics, the most widely used method for diagnosing malaria remains the manual counting of parasites within infected red blood cells (RBCs) from microscopic inspection of Giemsa-stained blood films (2); a method that has remained largely unchanged in nearly 120 years (3). Besides diagnosing malaria, Giemsa staining is also the cornerstone of laboratory research that involves parasite tissue culture (4), including drug and vaccine efficacy trials. However, the identification and counting of parasites is a time-consuming process that requires trained microscopy technicians (5). Moreover, manual evaluation can be erroneous and vary between slide readers (6–8).

Recent advances in machine learning (ML) have provided an opportunity to explore automating the detection of parasites from cytological smears (9). Generally, these efforts focus on some or all of a sequence of computational tasks: (a) cell segmentation (the partitioning a digital image into multiple segments such as pixels) and the detection of individual RBCs, (b) parasite identification and discrimination between infected and uninfected cells, and (c) subclassification of the different stages of parasite development. Early efforts mostly applied methods based on histogram thresholding and morphological operations, extracting hand-crafted features, and classification using traditional ML methods (10–15). Recently, there has been a general trend towards using deep learning methods for feature computation (16–20) as well as cell segmentation (21–23).

Despite significant improvements in model performance, however, there is currently no widely used tool for the automated detection and staging of malaria parasites. Making models available to the wider community would enable their use in routine malaria research, where standards of parasite nomenclature and parasitaemia count vary between users and groups and provide a route to field testing to advance the automated clinical diagnosis of malaria from smears. A major caveat that has held back automated approaches to date is the absence of standardisation in the preparation of thin blood films, which results in extensive staining and lighting variations between laboratories (24). Moreover, introducing a usable method requires a processing pipeline from the raw image to result, with only a few studies focussed on all aspects of the pipeline. For example, most studies have developed methods for datasets of images of single segmented RBCs (20). This method requires manual cell segmentation by a microscopist and neglects the effect of dust, debris, staining artefacts, or neighbouring cells on parasite detection. Only a few studies have also combined parasite detection with the classification of the different stages of the intraerythrocytic development cycle (IDC). Furthermore, treating parasite development as a classification problem disregards information on progression within and between the individual stages; progression through the IDC is a continuous process and experts disagree on the boundaries between the different life stages (10, 21, 25, 26). Automation has the potential to save time and add to number of RBCs sampled for both diagnosis and lab usage, however, its usage in diagnosis would require confidence in the analysis process; if results are accessible for review post-analysis by a microscopist, such a system is more likely to be implemented as a robust decision support tool.

Here, we present a machine learning method that combines cell segmentation, parasite detection, and staging in *Plasmodium falciparum*. We use a combination of residual neural networks (ResNets) with transfer learning to quantify parasitaemia and categorise IDC stages in both an accurate and standardised manner. Moreover, we present the first application of a regression model to order life stage categorisation that accurately recapitulates the progression of the IDC. Bringing these approaches together, we have built a web tool prototype, called PlasmoCount, which allows for versatile detection and stage categorisation of malaria parasites from Giemsa-stained images with an interactive assessment of the results. We test the performance of the tool with independent test sets and observe high performance across all models. We demonstrate that the web tool can be used in conjunction with mobile devices from image capture to assessment, opening up new opportunities for rapid, low-cost automated diagnosis from gold-standard smears. We believe this approach can bring new rigour to *Plasmodium* biology studies based on the inspection of smears and as a platform for on-the-go support in clinical settings where Giemsa-based cytological smears are used as the primary tool for the diagnosis of malaria.

## Methods

### Sample preparation

An image dataset was collected by each research center according to the protocol as follows without further instructions to allow for experimental variation. Varied *P. falciparum* parasites (strains 3D7, NF54, DD2 and D10 depending on the research group) were cultured with a parasitaemia of around 5% according to standard protocols in the individual laboratories. Thin blood films were prepared and air-dried for 1-2 minutes. Smears were then fixed in methanol for 30 seconds and stained by flooding the slide for 15 minutes in a fresh Giemsa solution of 10% Giemsa stain in a phosphate-buffered solution. The smear was then washed in water and air dried. Slides were imaged under 100% oil immersion with a 100x objective and saved in TIFF format. For phone capture, images were taken as JPEG files with 2x optical zoom on an iPhone 8 by manually aligning the phone camera with the microscope eyepiece. Details on sample and imaging specifications from each dataset can be found in <u>Supplementary Table S1</u>.

### Labelling pipeline

We used a model-assisted approach for labelling using the LabelBox platform (27) as this greatly improved labelling speed and performance. Each labelling round contained ∼100 raw images of Giemsa smears. Predictions from the RBC detection model and malaria identification classifiers were uploaded as pre-labels using the LabelBox Python SDK. Annotators could correct, add, and delete bounding boxes around each RBC and choose from three labels: *infected, uninfected, unsure*. At this stage, each image was corrected only by one annotator. RBCs labelled as either *infected* or *unsure* were automatically uploaded to a second labelling round as crops alongside the original image. Annotators could choose one label from *ring, trophozoite, schizont, gametocyte, uninfected cell, not a cell (e*.*g. debris, merozoites, etc*.*), multiple infections*, or *unsure*. RBCs labelled as *ring, trophozoite*, or *schizont* were resubmitted for a third labelling round where annotators could choose multiple labels: *early ring, late ring, early trophozoite, late trophozoite, early schizont, late schizont, multiple infections, unsure*. For both labelling rounds, each RBC was labelled by three different annotators using random allocation. All images were labelled five times by five designated annotators from three different research centers for the test set.

### Data analysis

A ResNet-50 architecture (28) pre-trained on ImageNet (29) was used for the backbone of the Faster R-CNN object detection model and all classification models. For regression of the life stage development, we adopted a smaller Resnet-34 model (28) pre-trained on ImageNet to avoid overfitting by extending the model with two fully-connected (FC) layers (1024, 512; ReLu activated) preceded with batch normalisation and dropout (p=0.5) and a final FC layer (16) that provides the life stage measure. Training data was split into training and validation sets by 80/20 with random sampling. The validation set was used during the training phase to set the learning rate and determine early stopping. Data augmentation methods were used that included dihedral transformations, rotation, flipping, and random lighting changes to increase the variety of our training data. For the Faster R-CNN model, we used stochastic gradient descent for optimisation and a learning rate of 0.01 with step-wise learning rate decay. For all other models, we used the Adam optimisation algorithm (30) and adopted a one-cycle learning rate policy (31).

### Web application architecture

PlasmoCount is a prototype available on www.plasmocount.org (password for access available on request). The web server was built on a Flask framework and provides application programming interface endpoints to connect the front-end interface to the ML models. The front-end was developed using the React web application framework and the Plotly interactive graphing library. The results are stored in cloud storage using Google Storage. When a user uploads their images, a unique job gets created, data is submitted to the backend, and the user is redirected to the job-associated results page. The user also receives an email at the email address provided with a link to this page for future reference. The Flask backend listens to updates in the cloud storage; once the analysis is completed, the results get returned to the front-end where they are visualised with interactive components using React and Plotly.

## Results

### Data

Given the inherent inconsistencies of manual inspection, counting, and staging of malaria parasites by Giemsa-stained cytological smear, we sought to develop a machine-learning based approach that could automate analysis of standard laboratory-acquired images of *Plasmodium* cultures. Six datasets of microscopy images of thin blood films of *P. falciparum* cultures were generated using a standardised Giemsa protocol described in the <u>Methods</u> section. Each dataset was collected by a different research centre with one dataset left out for testing (<u>Fig. 1A</u>; <u>Supplementary Table S1</u>). Datasets were labelled in five labelling rounds by ten expert malaria researchers, familiar with Giemsa-smears, across two different laboratories. Labelling was split into two steps: marking out bounding boxes around uninfected and infected RBCs and subsequent life stages classification.

**Figure 1.**
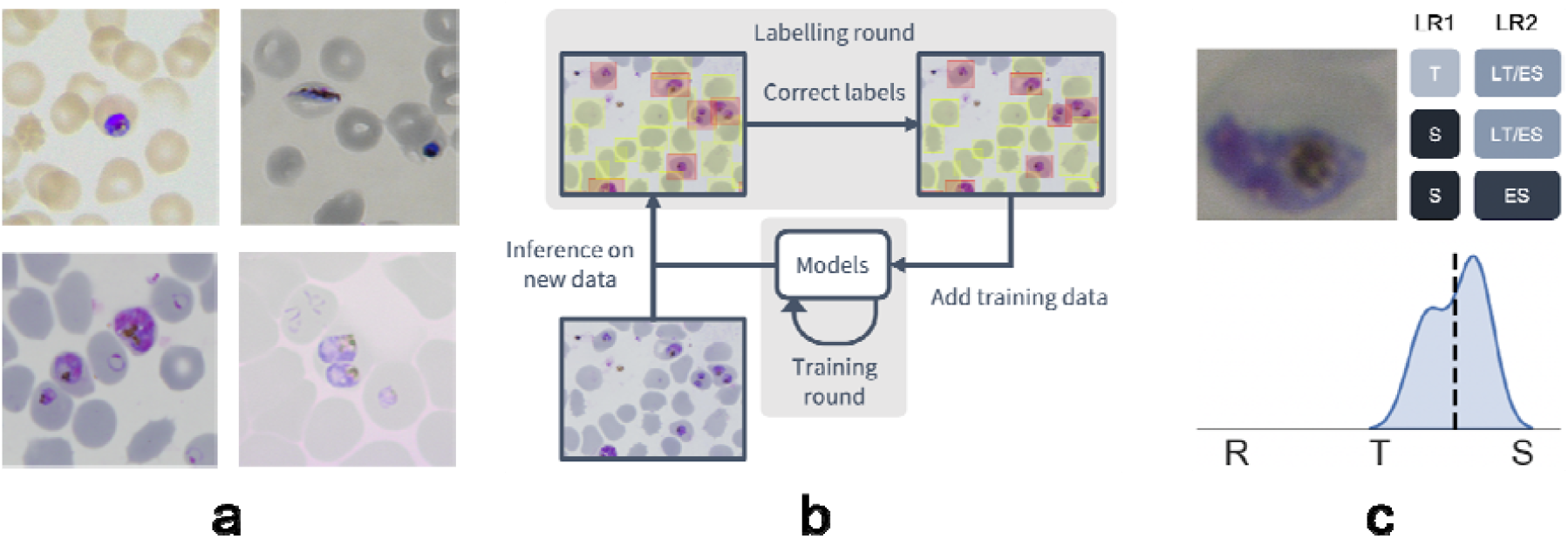
Data collection workflow. **a**. Example images of the *P. falciparum* dataset. Datasets were collected by six different research centers resulting in staining and lightning variations. **b**. Model-assisted labelling workflow. At each labelling round, model predictions were imported and loaded as editable annotations on an image. These annotations were corrected by annotators and added to the training data for a new training round. This process was then repeated with the retrained object detection and malaria classification models. **c**. Example of disagreement between annotators on parasite stage classification (R, Ring; T, Trophozoite; S, Schizont). Whereas the first labelling round (LR1) only captures the disagreement between two stages, the second round (LR2) reveals that this is due to the parasite existing between these two stages (LT, Late Trophozoite; ES, Early Schizont). The final value for the image is calculated by averaging labels across all annotators.

To facilitate bounding box demarcation, we used a model-assisted labelling workflow via the LabelBox platform (27). A new dataset was made available with bounding boxes drawn on uninfected and infected RBCs in images by the model after every training round. For the first labelling round, we trained on a dataset of *P. vivax* infected blood smears from the Broad Bioimage Benchmark Collection (32). Annotators were then asked to correct model predictions made on the *P. falciparum* dataset (<u>Fig. 1B</u>). According to standard IDC asexual stages (ring/immature trophozoite, trophozoite/mature trophozoite, schizont) and gametocytes, infected RBCs were next annotated by three different annotators per RBC. The first three stages correspond approximately to 0-12 hours post infection (hpi), 12-36 hpi and 36-48 hpi for asexual development, while gametocytes correspond to sexual development. Cells could also be labelled as multiple infections (more than one parasite per infected RBC) or marked as “*unsure*”. Whilst comprehensive, this classification task resulted in only 60% unanimous agreement between <u>all</u> annotators, with most disagreements occurring at the boundaries between IDC stages, particularly between trophozoite and schizont stages. Annotators never labelled the same image twice, so no data was collected on intra-annotator consistency; however, 9% of the collected labels were “*unsure*” unveiling intra-annotator uncertainty. Life stage assessment was repeated a second time for IDC stages but this time allowing annotators to select multiple values to account for uncertainty at boundaries. In addition, each IDC stage was subdivided into early and late stages (<u>Fig. 1C</u>). For example, a parasite between the ring and trophozoite stages could be annotated by selecting both “*late ring*” and “*early trophozoite*”. To provide a numerical score associated with labels, classifications were converted to a numeric scale (ring=1, trophozoite=2, schizont=3) and averaged across all annotators (<u>Supplementary Figure S1</u>). Multiple infections or cases where the annotators were unsure were discarded. In total, this led to a dataset with 38,000 different RBCs of which 6% were infected.

### Red blood cell detection

Having developed a standardised data set, we next trained a Faster R-CNN object detection model (33) to detect both uninfected and infected RBCs in microscopy images of thin blood smears. Faster R-CNN is a state-of-the-art object detection network that consists of a region proposal network (RPN) and a Fast R-CNN network that classifies the proposed regions (34). This approach has recently shown some promising results in cell segmentation in blood smears of *P. vivax* (23). We used a Faster R-CNN model pre-trained on the COCO dataset (35), fine-tuned on our *P. falciparum* dataset but also trained on the aforementioned *P. vivax* data. The classification task was not included at this first stage, instead focussing only on cell segmentation, aiming to make the model robust to variations between laboratories by applying multiple data augmentation methods. These methods included rotations, flips, blur, and colour shifts. False positives in the form of debris, merozoites (generally smaller than RBCs) or leukocytes (generally larger than RBCs) were excluded by applying a cut-off based on the area distribution of the detected bounding boxes with those 4 standard deviations above the mean area across the image discarded. Finally, cells that touched the border of the crop were excluded. A non-maximum suppression was applied using an Intersection Over Union (IOU) threshold of 0.5 to exclude overlapping bounding boxes. The Average Precision (AP) at an IOU threshold of 0.5 was 0.98 on the *P. vivax* test set and 0.99 on our test *P. falciparum* set (<u>Fig. 2</u>). Thus, we can extract individual RBCs from the background effectively as an input for further classification.

**Figure 2.**
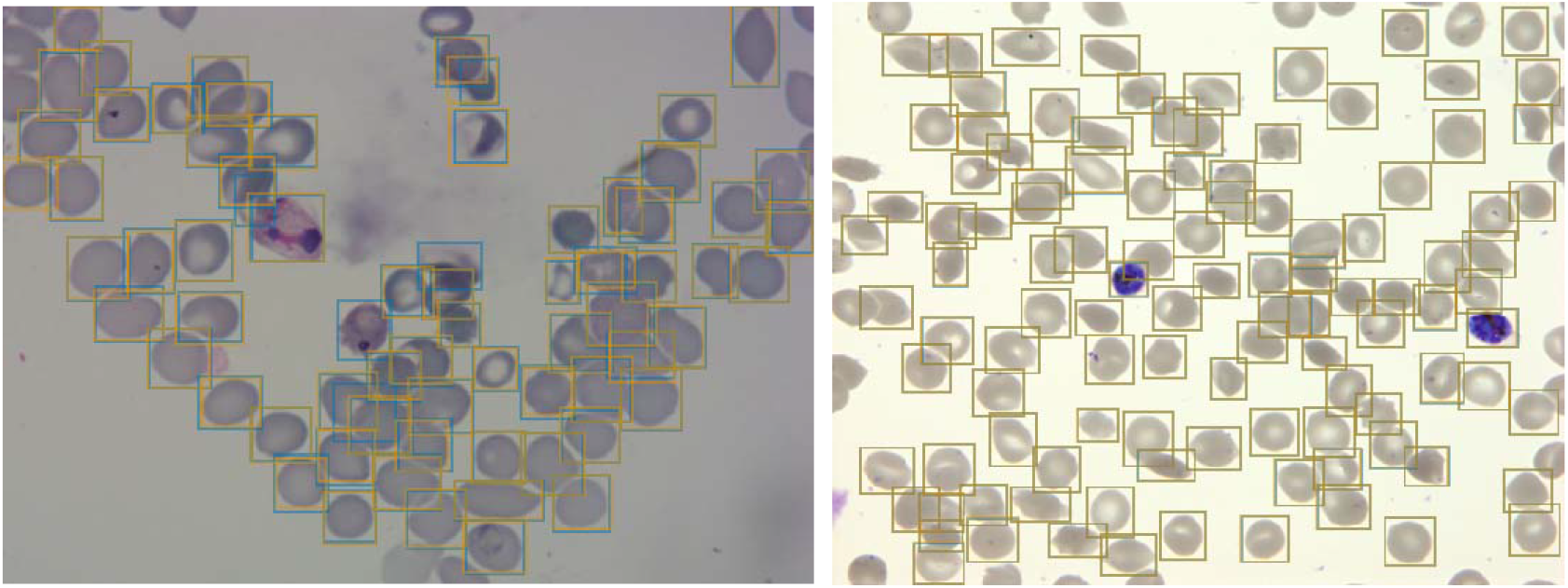
Object detection of red blood cells on thin smear images. Ground truth labels are shown in orange, predictions shown in blue. Examples shown for *P. vivax* (left) and *P. falciparum* (right) test data. The object detection model detects individual red blood cells with high precision despite the presence of dust, residual stain, and overlap with other cells.

### Infected and uninfected red blood cell differentiation

Having shown our ability to isolate RBCs with high precision, we next turned our attention to the delineation of infected versus uninfected cells. RBCs were cropped and scaled to 70×70 pixel resolution. We trained a Resnet-50 architecture (28) pre-trained on the ImageNet dataset (29) to classify infected RBCs using similar data augmentation methods to those described for the object detection model; however, colour augmentations were reduced in order to preserve colour properties of parasites inherent to Giemsa staining. To address the heavy class imbalance, training data was undersampled at each epoch (the number of times that the learning algorithm works through the entire training dataset) to generate a 1:1 ratio of infected:uninfected RBCs. Using this approach, we obtained an accuracy of 0.998 with an area under the receiver operating characteristic curve (AUC) of 0.979 on the test set with only images where all annotators agreed included (<u>Table 1</u>). Most misclassifications observed were false positives due to debris on top of an uninfected RBC or the presence of neighbouring infected cells. We found that annotators agreed only on 69% of images with a relative difference in parasitaemia of up to 5% (<u>Supplementary Figure S2</u>). Moreover, compared to cases where all other annotators agreed, three annotators achieved accuracies of 0.998, two of 0.999, and one of 1.000; demonstrating that our model’s error is indistinguishable from the error of an annotator.

**Table 1.**
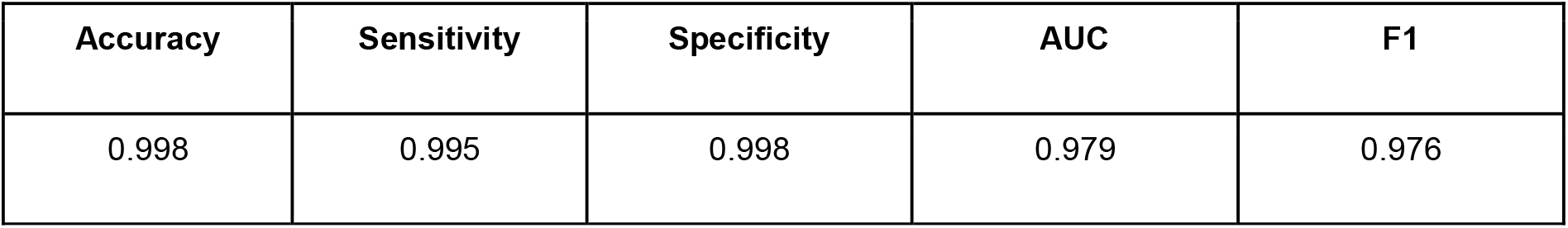
Performance of Resnet-50 model on malaria detection in images of segmented red blood cells. Metrics were calculated only for images on which all annotators agreed.

| Accuracy | Sensitivity | Specificity | AUC | F1 |
| --- | --- | --- | --- | --- |
| 0.998 | 0.995 | 0.998 | 0.979 | 0.976 |

### Life cycle stage classification

Having established a model for the classification of infected versus uninfected RBCs in cropped images, we next sought to further classify infected cells into the different parasite IDC stages and gametocytes. Rather than expanding our existing classifier to include the stages as discrete classes, we instead approached progression through the IDC as a continuous process where the stage exists on a numeric scale (ring=1, trophozoite=2, schizont=3). To achieve this, we adapted a pre-trained Resnet-34 classifier (28) to a regression model. We pooled and flattened the final convolutional layer using average and maximum pooling and extended the model with two fully-connected neural network layers of 1024 and 512 neurons with ReLU activation. Each of these layers is preceded by batch normalisation and dropout with a probability of 0.5. The output of the 512 neurons fully-connected layer feeds into a final fully-connected layer of 16 neurons to measure the life stage. Using this model, we achieved a root mean squared error (RMSE) of 0.23 in the test set, with most errors observed between late trophozoite stages and early schizont stages (<u>Figure 3A</u>). High levels of disagreement were found between annotators (<u>Figure 3A</u>; <u>Supplementary Figure S3</u>), and most of the model predictions generally fell within these label boundaries. Moreover, the model provided an ordering of the infected RBCs following the IDC (<u>Figure 3B</u>). Thus, we are able to describe the IDC with more detail than with a three-class classification approach.

**Figure 3.**
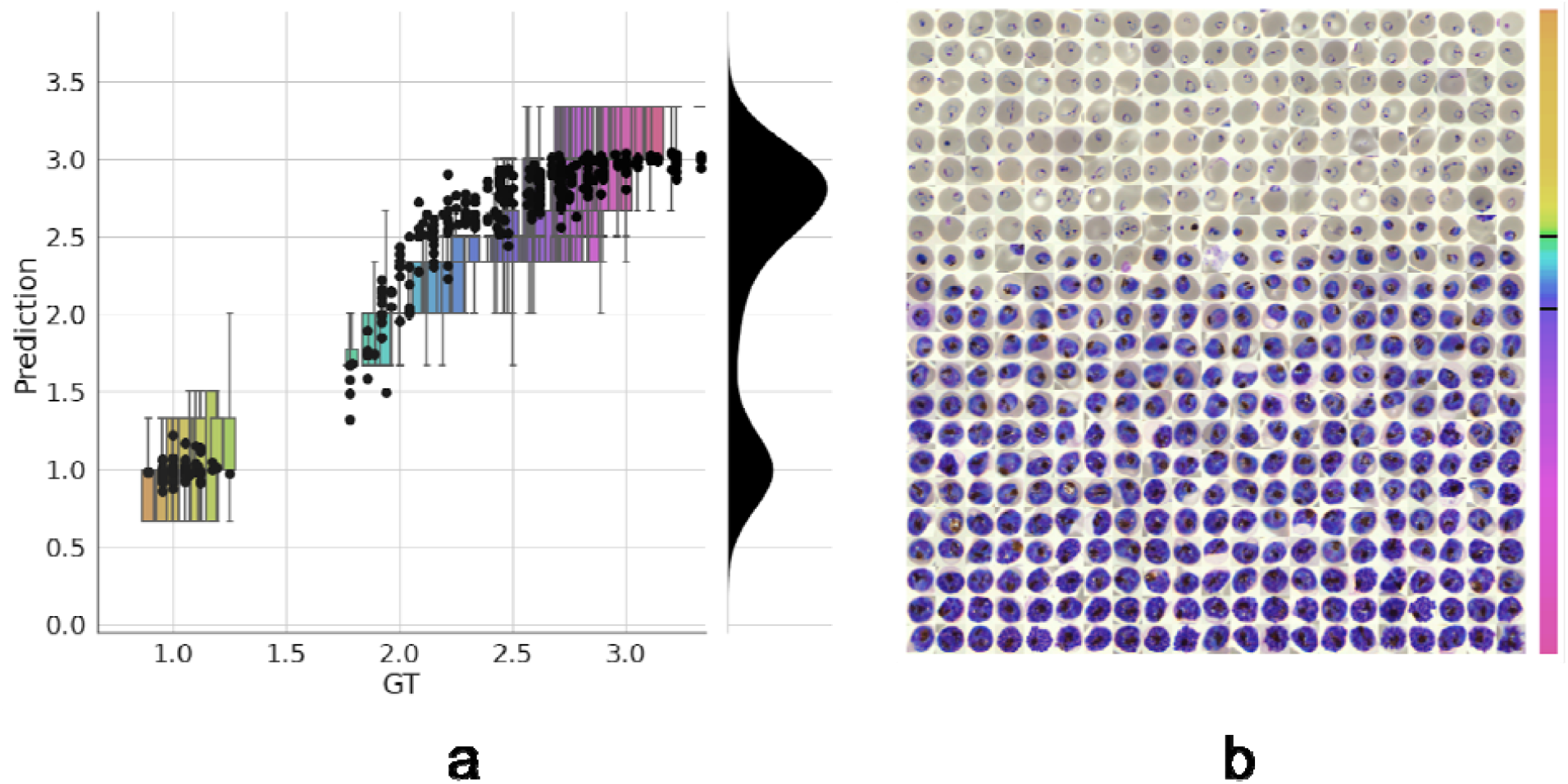
Model prediction on parasite intra-erythrocytic cycle (IDC) development. **a**. Model predictions are marked with a black dot. Labels were converted to a numeric scale (ring=1, trophozoite=2, schizont=3) and averaged across all annotators to set a ground truth (GT) label. Boxplots show the label distribution across annotators with error bars determined by the outermost data values. Density plot shows the predicted life stage distribution within the sample. Colors represent progression through the IDC as defined by the GT. **b**. After learning from the averaged GT labels, the model successfully orders all detected infected RBCs in the independent test set based on its intra-erythrocytic cycle (IDC) life stage predictions (left to right; top to bottom). Colorbar represents progression through the IDC as predicted by the model. Top and bottom black lines represent the arbitrary cut-off points used by PlasmoCount between the ring and trophozoite (cut-off=1.5) and trophozoite and schizont (cut-off=2.5) stages, respectively.

**Figure 4.**
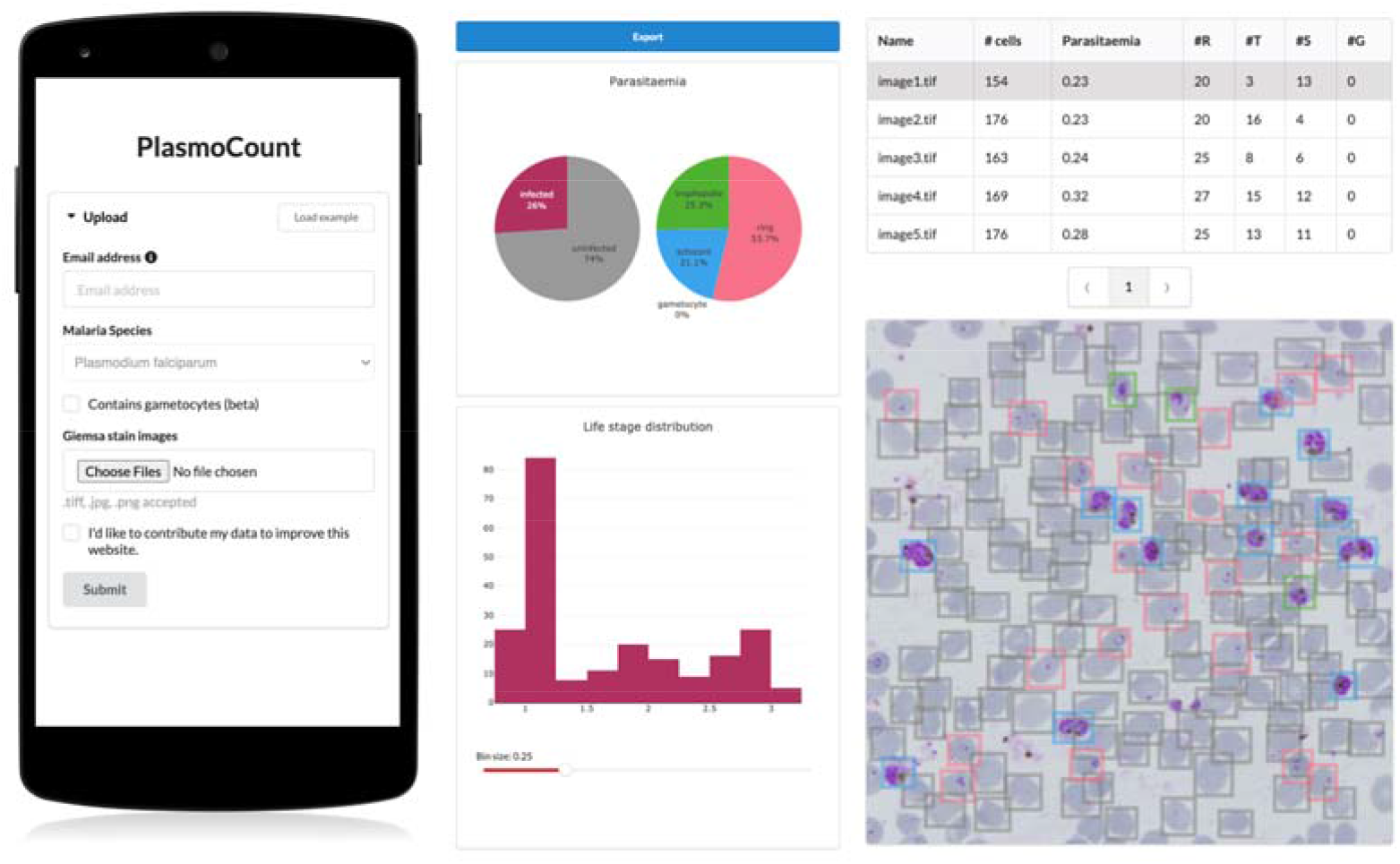
PlasmoCount, a mobile friendly tool for automated assessment of Giemsa-stained cytological smears. In the upload form, users can attach their images of Giemsa-stained thin blood films. The client sends the data to the server to be passed to the models; the results then get sent back and are displayed in a summary section and a table. The summary section is divided into parasitaemia pie charts and a histogram of the IDC life stage distribution. The rows in the table correspond to individual images and are selectable; this will display the corresponding image with overlaying model predictions. The user interface is optimised for mobile phone dimensions, allowing users to use a mobile device for both image acquisition and data analysis. Every job has a unique ID associated with it; this allows users to come back to their results from multiple devices at any time.

Gametocytes form when asexual parasites commit to sexual development and follow a morphological progression different from the IDC stages. They cause no clinical manifestations and are generally cultured *in vitro* specifically to study parasite transmission; in line with this, our test set did not contain any gametocytes. However, to show that our method could be extended to include the sexual cycle of the parasite, we trained a Resnet-50 classifier to discriminate between RBCs infected with asexual and mature gametocytes and assessed performance with 5-fold cross-validation. For this purpose, we included a dataset in our training set that was optimised for gametocyte formation, containing a mixture of sexual and asexual parasites. Because of the imbalance between these classes in the training set, gametocytes were oversampled to obtain a 1:1 ratio. This resulted in a mean (± standard deviation) accuracy of 0.991 ± 0.007 and AUC of 0.971 ± 0.021; showing that we can discriminate between asexual and sexual parasites preceding further discrimination within each developmental cycle.

### PlasmoCount: A web tool for automated parasite detection and staging

Having demonstrated high performance across models on an independent test set of Giemsa-stained thin blood films, we next sought to generate a user interface that would allow our ML approach to be applied in different lab settings. Towards this, we developed PlasmoCount: an online web tool that can take as input multiple raw images of a Giemsa-stained thin blood film and outputs measures of parasitaemia and parasite life stage development. The images are run through the object detection model; the resulting RBCs are run through the malaria detection and staging models sequentially. The output contains two interactive sections: a summary section that reports parasitaemia and the IDC stage distribution in the sample and a table with results on the individual images. The interactive component within the summary section allows users to change the histogram bin size of the histogram and click on the individual bins to display the corresponding RBCs for interpretation of the life stage discrimination. The table describes, for each image, the number of RBCs, parasitaemia percentage, and the number and fractions of the different IDC and gametocyte stages of the parasite and data can be exported in CSV format. Moreover, the user can click on the individual rows to display the model predictions for each image as a measure of quality assurance. The web tool has been optimised for mobile phones in conjunction with images directly taken with the phone camera or a cell-phone microscope system. Beta-testing with users from different labs, testing for accuracy and utility, was found to perform well with a mobile phone looking down the microscope even without an adaptor (<u>Supplementary Figure S4</u>). Combining the power of the ML model approach with the utility of a mobile interface-based web tool demonstrates the power of machine learning to identify and classify parasites and its ability to rapidly assist in the assessment of parasite cultures via cytological smears.

## Discussion

Here, we present a machine learning approach and user-orientated web tool for the detection and staging of *P. falciparum* from Giemsa-smeared cytological smears. Using this simple approach, we obtain state-of-the-art performance in cell segmentation and detect parasites with high performance on an independent test set. Our object detection model can detect RBCs with an average precision (AP) of 0.99 on our *P. falciparum* test set and an AP of 0.98 on a *P. vivax* dataset previously tested with the novel Keras-RCNN model (mAP=0.78) (23) and a CellProfiler pipeline (mAP=0.61) (23, 36) at an IOU threshold of 0.5. Moreover, our classification model of infected RBCs reaches a near-perfect accuracy with a sensitivity of 0.995 and specificity of 0.998, respectively; this is comparable to state-of-the-art methods for the classification of segmented RBCs (20, 37, 38). Indeed, we could further improve our method by replacing our object detection method with a Mask-RCNN model where each cell is overlaid with segmentation masks rather than bounding boxes. This approach has been successfully applied in nucleus segmentation (39, 40) and would reduce the confounding of neighbouring cells on parasite detection. Lastly, we have introduced a staging model that recapitulates IDC progression of the parasite and allows for more refined interpretation when reading slides. Recent studies have aimed to enhance IDC life stage classification by adding early, mid, and late sub-stages (25). Using a regression model, we are able to differentiate the IDC stage of the parasite more precisely and prevent penalising predictions that have been classified as a different sub-stage of the parasite but are developmentally not far removed.

We have shown that whereas experts disagree on parasite stages, using an automated approach to malaria detection enables robust standardisation in the evaluation of Giemsa smears, providing quantitative measures, for example, when applied to high-throughput experiments. We have defined the ground truth as a summary of labels from multiple annotators and laboratories. Using this method, our model is able to generally order the developmental stages of individual cells and recapitulate the IDC life cycle of *P. falciparum*. However, the ground truth could be further refined by including more annotators and allowing annotators to select from a wider range of stages. Moreover, our pipeline could be supplemented by asking annotators to order cells as part of the labelling process as this has been shown to improve consensus among annotators in fluorescent imaging data of *P. falciparum* (26). It should be noted that there is as yet no method of validating this ground truth; ultimately, it is defined by the experts who evaluate the slides, and this phenomenon has led groups to correct standard datasets (41). Methods will have to establish “gold standard” slide/image sets, not only for the assessment of reader competency but also to ensure a better benchmark for the design of automated methods.

In this work, we have focussed on discriminating between the IDC stages of the parasites as these asexually replicating stages cause symptoms in patients and are the main focus of clinical studies (42) as well as providing the source population for gametocyte investment (43). As a proof-of-concept, we have shown that we are also able to discriminate between asexual and mature sexual blood-stage parasites using ML methods. In the future, we would like to extend our regression approach to the sexual stages of the parasite, including progression through the five distinct stages of gametocyte development and differentiation into males and females. We have also not addressed cases where multiple parasites invade red blood cells, which is likely to confound the IDC stage prediction. Possible solutions include adding a local classifier for multiplicity, multilabel classification with the IDC stages, and object detection for detecting individual parasites.

Although we have developed PlasmoCount to detect *P. falciparum*, it is by no means limited to one species of *Plasmodium*. Preliminary testing suggests that the tool can also be applied to the study of round-the-clock blood samples from infections with rodent malaria parasites (*P. chabaudİadami*, clone DK), although this will need to be validated against a ground truth based on expert annotations (<u>Supplementary Figure S5</u>). Leukocytes are not typically present in our laboratory smears of *P. falciparum*, which generally use only processed RBCs from donated whole blood. We have, however, aimed to address the possibility of leukocyte presence by including a threshold based on the size distribution of cells in the sample but a more sensitive ML-based method could be added in the future. Such an approach will be fundamental as we aim to extend our method to the clinical diagnosis of human malaria where leukocytes, other components of whole blood (e.g. platelets) and even other pathogens may add further classes of non-RBC material to smears.

Automated methods can speed up the evaluation of thin blood smears; however, the bottleneck of image acquisition remains. In this study, we sought to provide a solution to the speed of acquisition by developing a tool that works in conjunction with mobile devices from image acquisition to assessment. If coupled with low-cost and portable microscope systems (44–46), this has the potential to increase screening throughput markedly and opens up opportunities for thin blood smears as a tool for qualitative diagnosis in the field where thick smears are preferred due to their higher sensitivity. Speed of image acquisition via a mobile device will generally be a lot faster than manual counting and recording of thin smears. There is also the possibility that PlasmoCount could be incorporated into higher throughput image acquisition system workflows (e.g. slide readers) to speed up accurate parasite quantification in a hospital/laboratory-based setting.

There are limitations to be considered especially in the use of ML methods in clinical practice. Although we have tested a dataset from independent research centers, measuring generalisation performance remains challenging due to technical differences between laboratories, and performance will heavily depend on data quality. Moreover, our method does not estimate uncertainty with its predictions which would improve the reliability of the results. To this end, we have developed a web tool that lets the user check their results for quality assurance and save them for traceability in the decision-making process. This also opens up an opportunity for continuous improvement of the models; by collecting feedback from the community, a human-in-the-loop labelling workflow could be implemented similar to our model-assisted approach.

Recent efforts have applied ML techniques to the detection of malaria parasites in microscopy images of cytological smears. The aim of this work was not only to further these developments but also to provide a tool that can be used by the malaria research community. Deep learning typically requires large amounts of data, which are not always widely available for medical applications due to the expert knowledge required. In this study, we have included data from six research centers and tested the tool’s functionality with various others. In this way, we enhance robustness across laboratories and aim to drive a community-wide effort to accelerate malaria research and ultimately adopt automated methods in a clinical diagnostic context.

## Data Availability

Original imaging data, experimental metadata and pseudolabels will be available from a publicly accessible Image Data Repository (pending upload and URL to be provided).

## Author contributions

MSD and JB designed the experiments; SY, JC, DW, PP, CU, MJ, MP, DA, MB, PG, JD, AO, and SR cultured parasites, stained and acquired images; MSD, GWA, MD and DMA developed the imaging workflow; MSD, MD and DMA optimised and carried out the machine learning workflow and data analysis. All authors contributed to writing the manuscript.

## Data and materials availability

Original imaging data, experimental metadata and pseudolabels will be available from a publicly accessible Image Data Repository (pending upload and URL *to be provided*).

