## Supplementary Information for "Automated detection and staging of malaria parasites from cytological smears using convolutional neural networks"

### **Affiliations:** ^1^ Department of Life Sciences, Imperial College London, London, UK. ^2^ Research Centre for Infectious Diseases, School of Biological Sciences, University of Adelaide, Adelaide, Australia. ^3^ Institute of Evolutionary Biology, and Institute of Immunology and Infection Research, School of Biological Sciences, University of Edinburgh, Edinburgh, UK. ^4^ Division of Infectious Diseases, Boston Children's Hospital, Boston, Massachusetts, USA and Department of Pediatrics, Harvard Medical School, Boston, Massachusetts, USA. ^5^ Mikrobiologisches Institut–Klinische Mikrobiologie, Immunologie und Hygiene, Universitätsklinikum Erlangen, Friedrich-Alexander-Universität (FAU) Erlangen-Nürnberg, Erlangen, Germany. ^6^ National Center for Genetic Engineering and Biotechnology (BIOTEC), Pathum Thani, Thailand. ^7^ QIMR Berghofer Medical Research Institute, Herston, QLD, Australia. ^8^ Burnet Institute, 85 Commercial Road, Melbourne, Victoria, Australia. ^9^ Google Research, Applied Science Team.

### **Short Title**: Machine learning for malaria parasite detection

Supplementary Information

Supplementary Figures S1-6

Supplementary Dataset S7

Supplementary Table

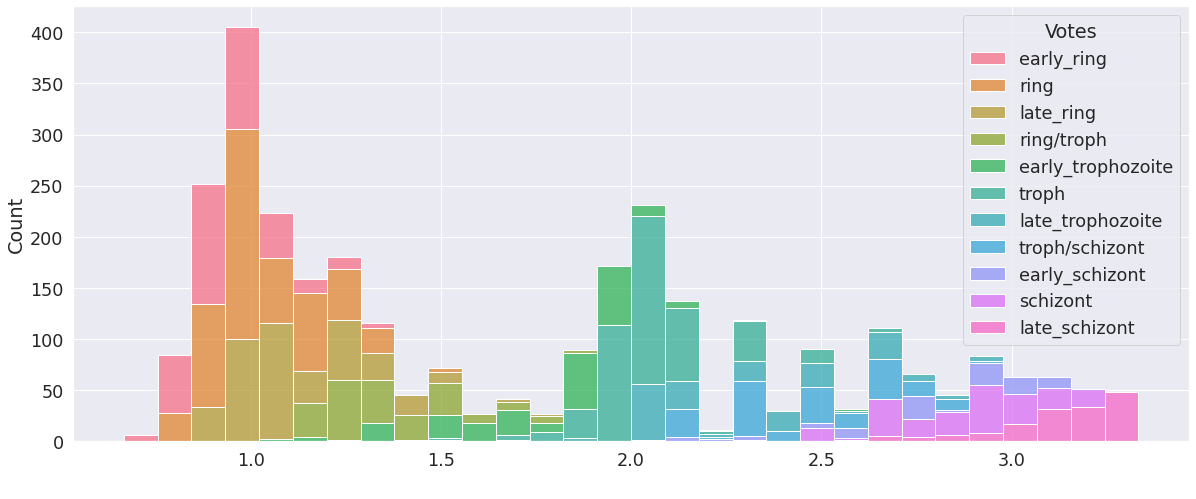

###### Supplementary Figure S1: Histogram of annotator labels for ground truth labels of intra-erythrocytic developmental cycle (IDC) stages. Annotators were shown an image of an RBC infected with a parasite from the IDC. Label options were early ring, late ring, early trophozoite, late trophozoite, early schizont, late schizont; a canonical ring would be annotated by selecting both “early ring” and “late ring”. Annotators were allowed to select multiple values. Labels were converted to a numeric with all labels assigned according to the scale of ring=1, trophozoite=2, schizont=3. All selected labels were merged and averaged to get a ground truth label, as presented on the x-axis. IDC stages are coloured according to individual labels from annotators.

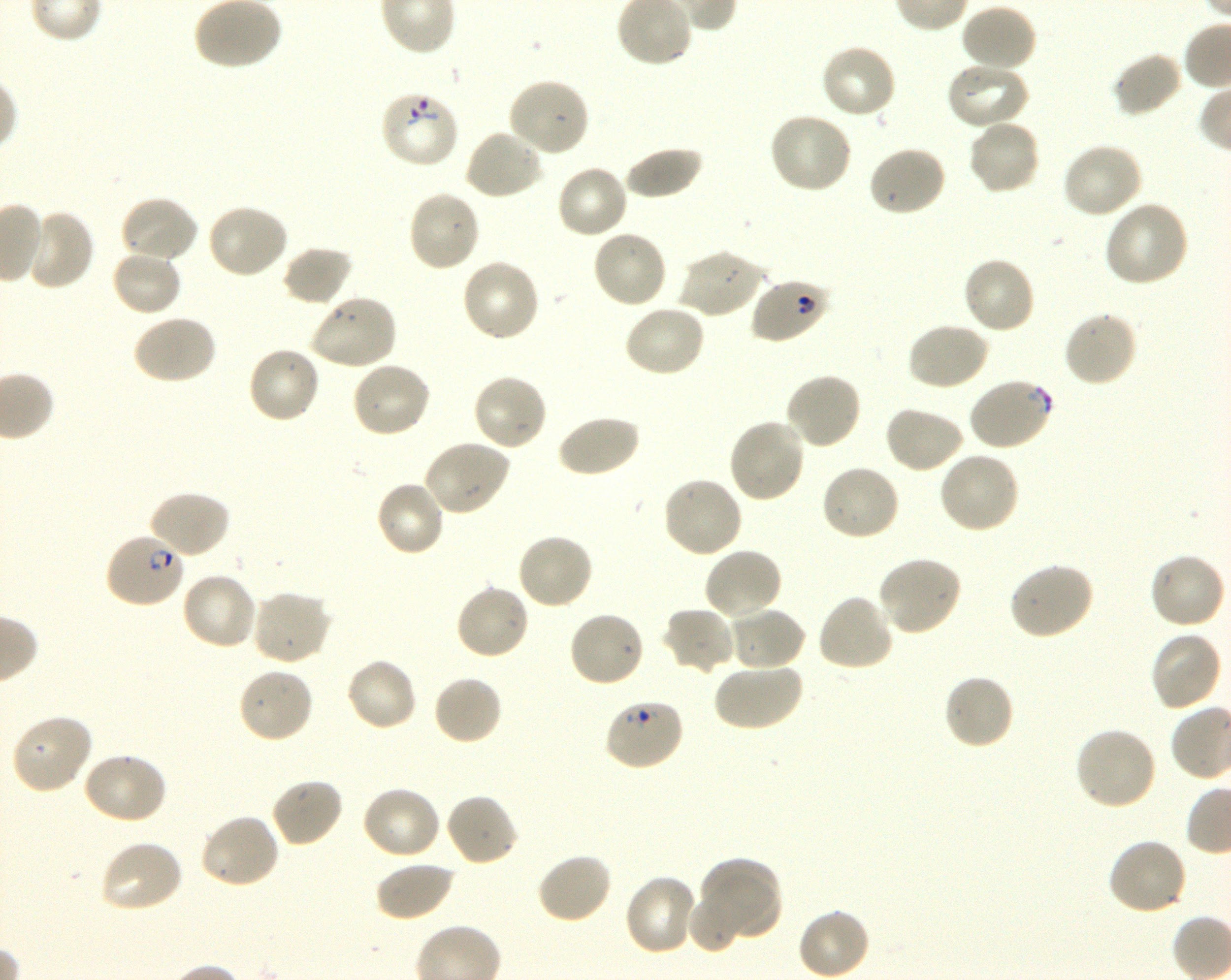

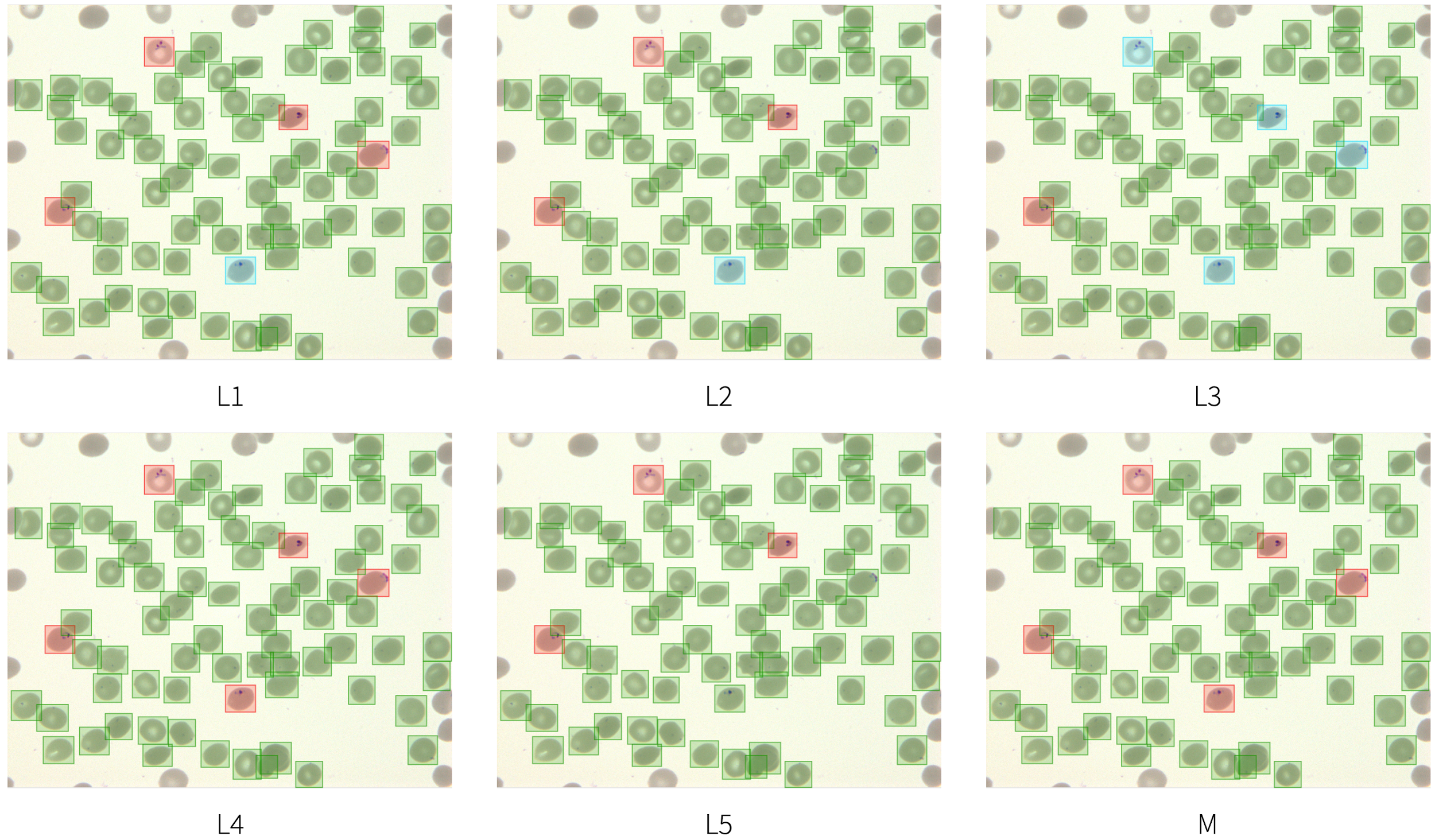

###### Supplementary Figure S2: Labelling variation in parasitaemia calculation. Five annotators (L1-5) across three different research centers were asked to correct bounding boxes around RBCs and label those infected. Only cells where all annotators agreed on infection were used for measuring performance. Uninfected cells (green), infected cells (red), and cells labelled as “unsure” (blue) are annotated. M=Model prediction.

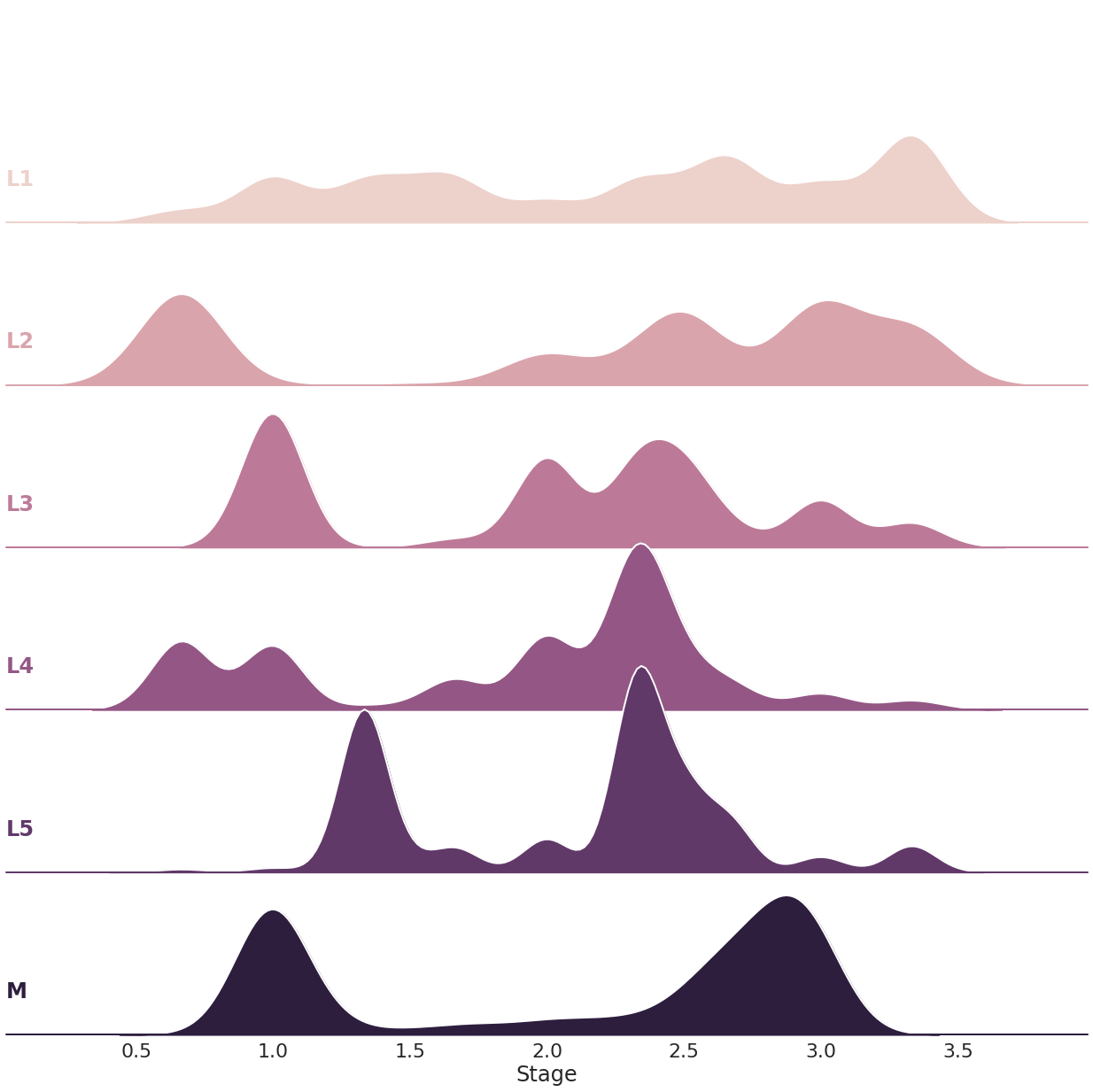

###### Supplementary Figure S3: Labelling variation in IDC life stage distribution. Five annotators (L1-5) across three different research centers were asked to label the same data set for segmented infected RBCs as a combination of early/late ring, trophozoite, and schizont labels. Labels were converted to a numeric scale where ring=1, trophozoite=2, schizont=3; e.g. a selection of late trophozoite and early schizont results in a value of 2.5. We observe large variability between annotators, highlighting the need for standardisation in the evaluation of blood smears and the potential of using automated methods towards this objective. M=Model prediction.

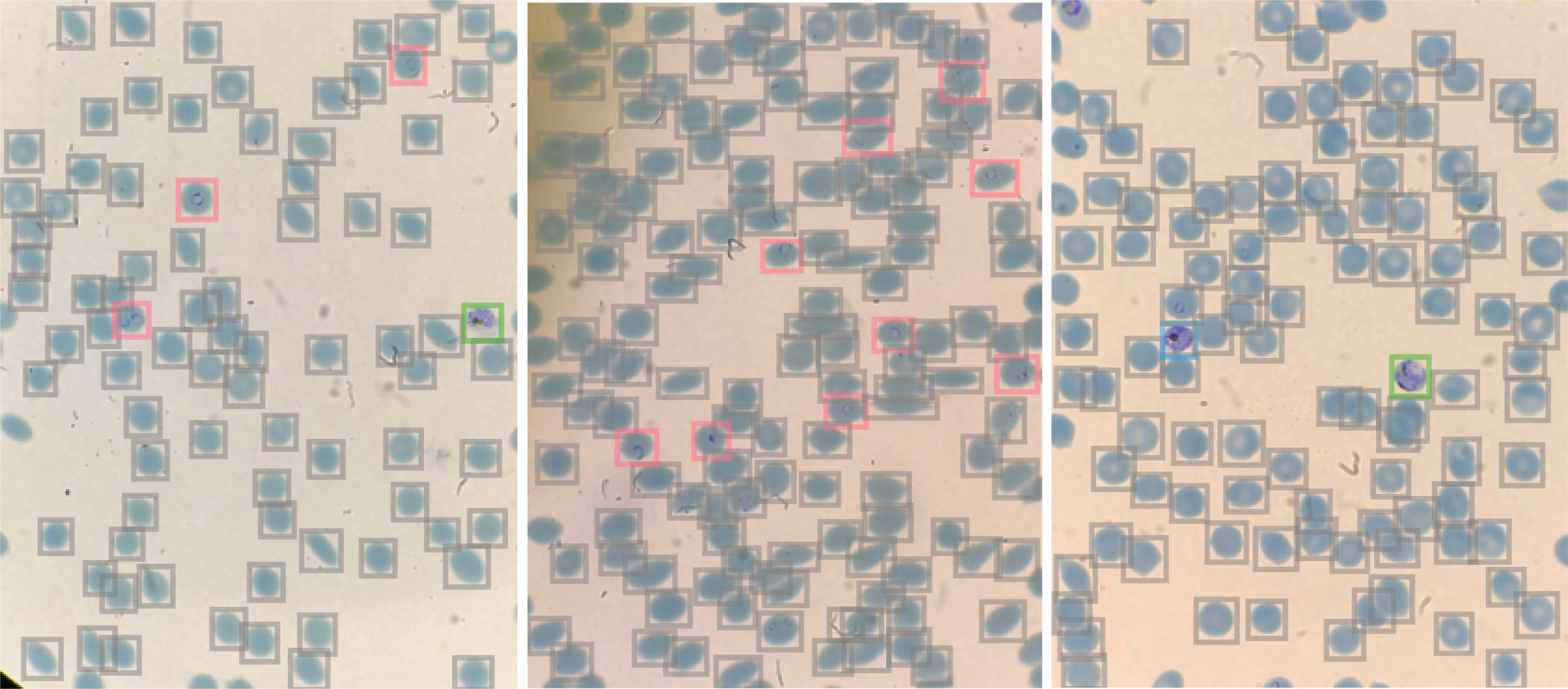

###### Supplementary Figure S4: PlasmoCount predictions using mobile phone pipeline. Images were captured with an iPhone 8+ camera at 2x zoom by aligning the camera with the microscope eyepiece (Nikon Ti2-E). Images were then uploaded to PlasmoCount for assessment of the blood films (ring=red, trophozoite=green, schizont=blue, uninfected RBC=grey).

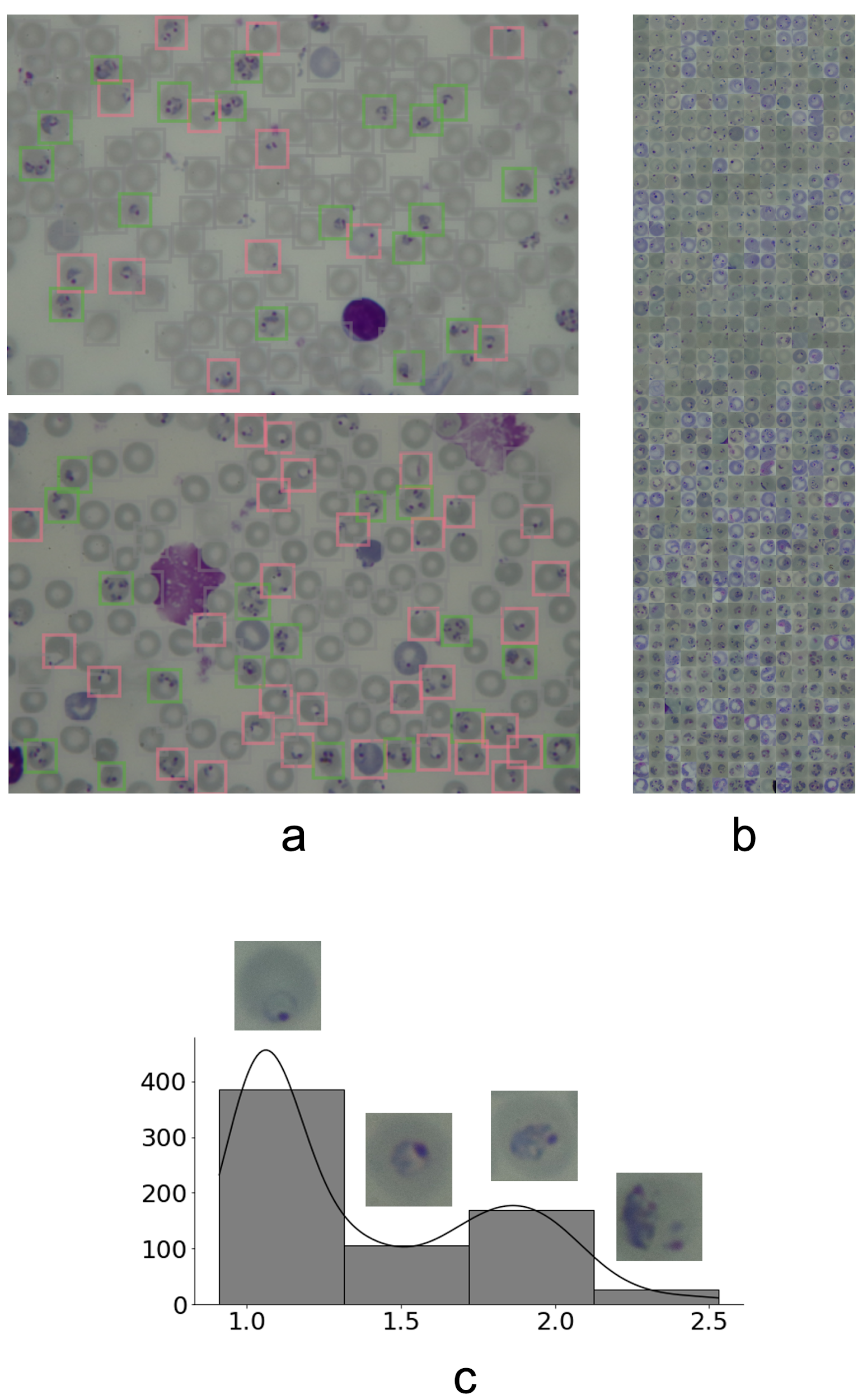

###### Supplementary Figure S5: PlasmoCount predictions on *P. chabaudi adami* clone DK data. Example images (a) show that using a per-image threshold for the area of bounding boxes can be sufficient to eliminate leukocytes and stain precipitation (ring=red, trophozoite=green, schizont=blue, uninfected RBC=grey). (b) Ordering of all detected infected RBCs by PlasmoCount (left to right; top to bottom). (c) Parasite distribution as predicted by PlasmoCount. Example RBCs are displayed for each bin. Bins shown generally correspond to ring, early trophozoite, mid trophozoite and late trophozoite stages according to our numeric scale (ring=1, trophozoite=2, schizont=3). A few gametocytes have been detected. These are not mistaken for rings and have been approximately placed in between mid trophozoites and schizonts. This suggests that future optimisation of the model may enable successful differentiation of IDC and gametocytes (which wasn’t attempted here) in other parasite species.

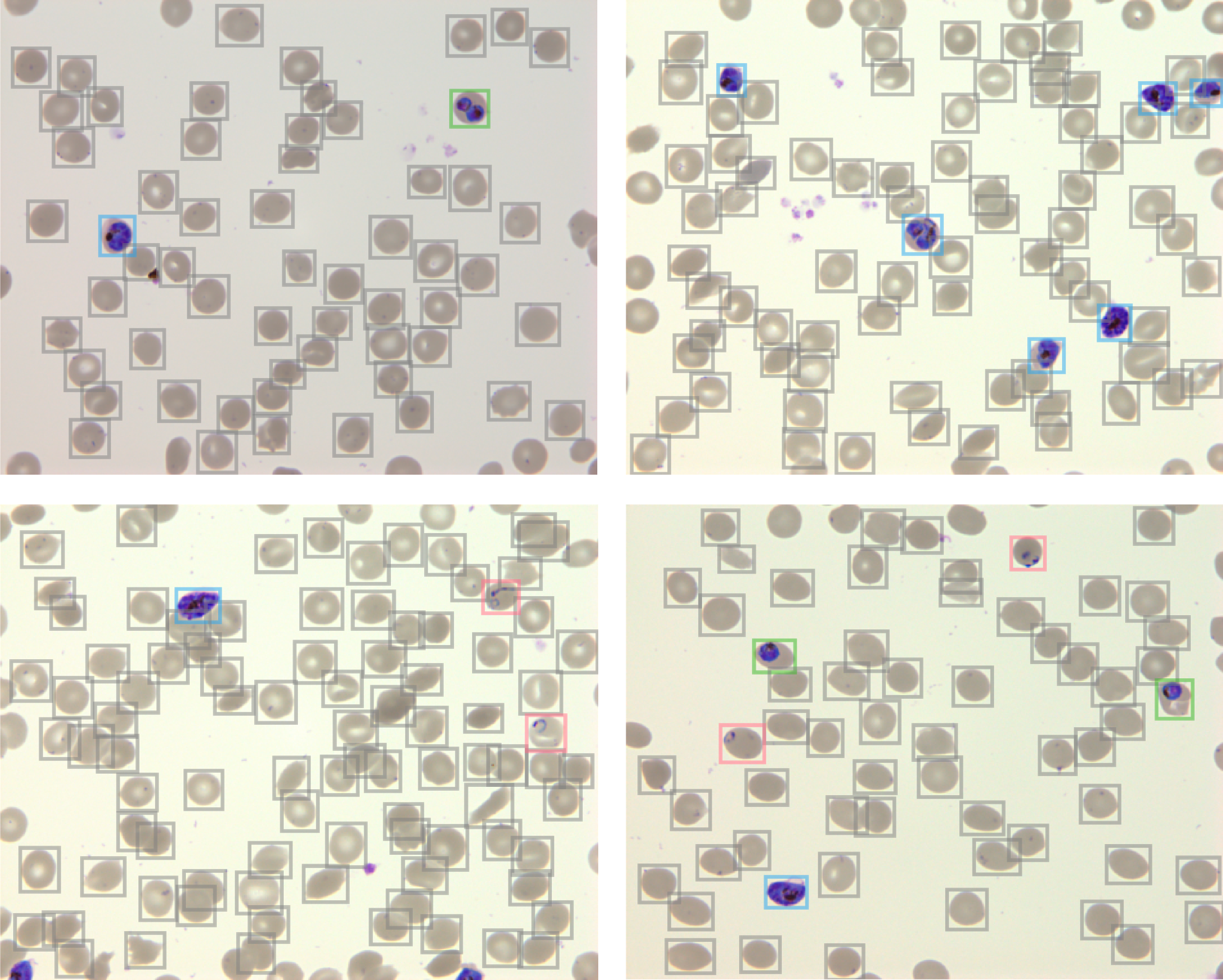

###### Supplementary Figure S6: PlasmoCount predictions on *P. falciparum* test set. Example images are provided as Supplementary Dataset (S7) for testing purposes on [www.plasmocount.org](http://www.plasmocount.org) (login details provided on request).

| **Microscope brand +**  **model** | **Lens objective (x)** | **Numerical aperture** | **Malaria species + strain** | **Parasitemia** | **Donor Blood Group** | **Cultivation** |
| --- | --- | --- | --- | --- | --- | --- |
| Olympus | 100 | 1.4 | *Plasmodium falciparum* 3D7 | 20% | O | Static culture |
| Zeiss Axioskop 40 | 100 | 1.25 | *Plasmodium falciparum* NF54 | 10% | A+ | Static culture |
| Leica DM750 | 100 | 1.25 | *Plasmodium falciparum* 3D7 | 4.4% | O+ | Static culture |
| Nikon Ti2-E Inverted Microscope | 100 | 1.45 | *Plasmodium falciparum* NF54 | >2% | A+/O+ | Static culture |
| Olympus LC20 | 100 | 1.25 | *Plasmodium falciparum* 3D7, DD2, D10 | 4-6% | O+ | Static culture |
| Olympus BX40 with INFINITY3-6UR camera | 100 | 1.30 | *Plasmodium falciparum 3D7* | 3% | O+ | Shaking culture |

###### Supplementary Table S1: Dataset specifications. Test set is highlighted.
