## Supplementary figures and images for "Automated detection and staging of malaria parasites from cytological smears using convolutional neural networks"

### 1.jpg

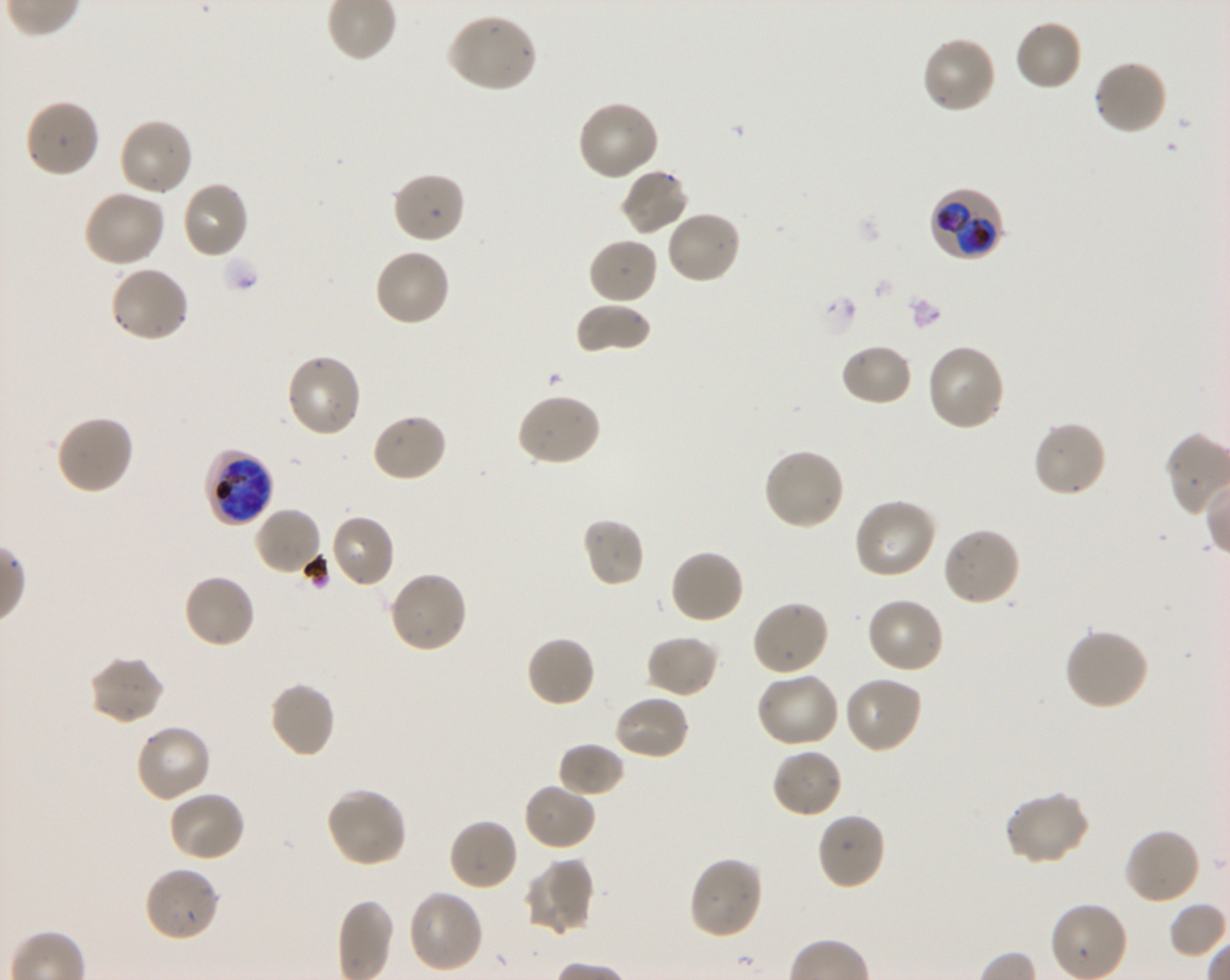

### 2.jpg

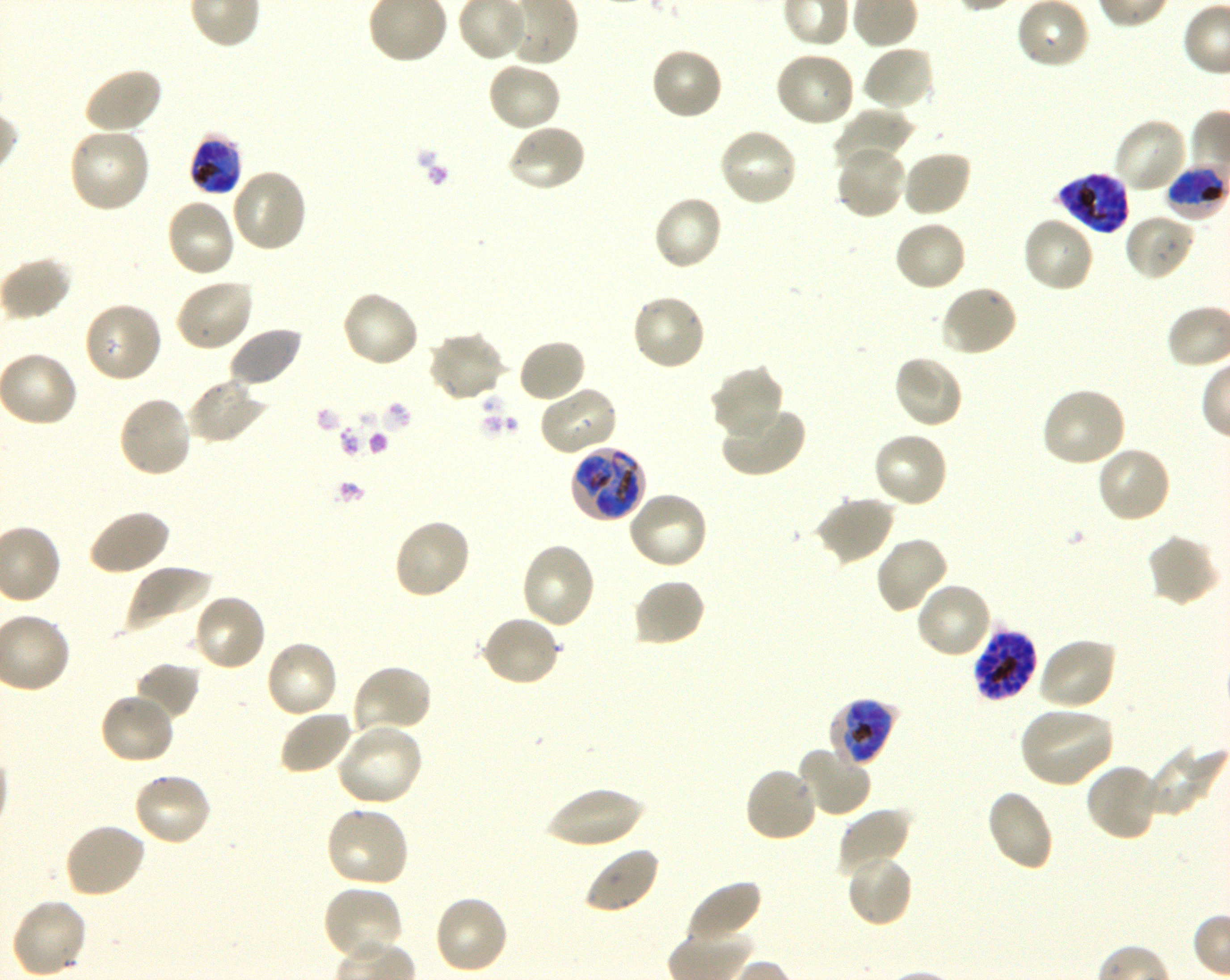

### 3.jpg

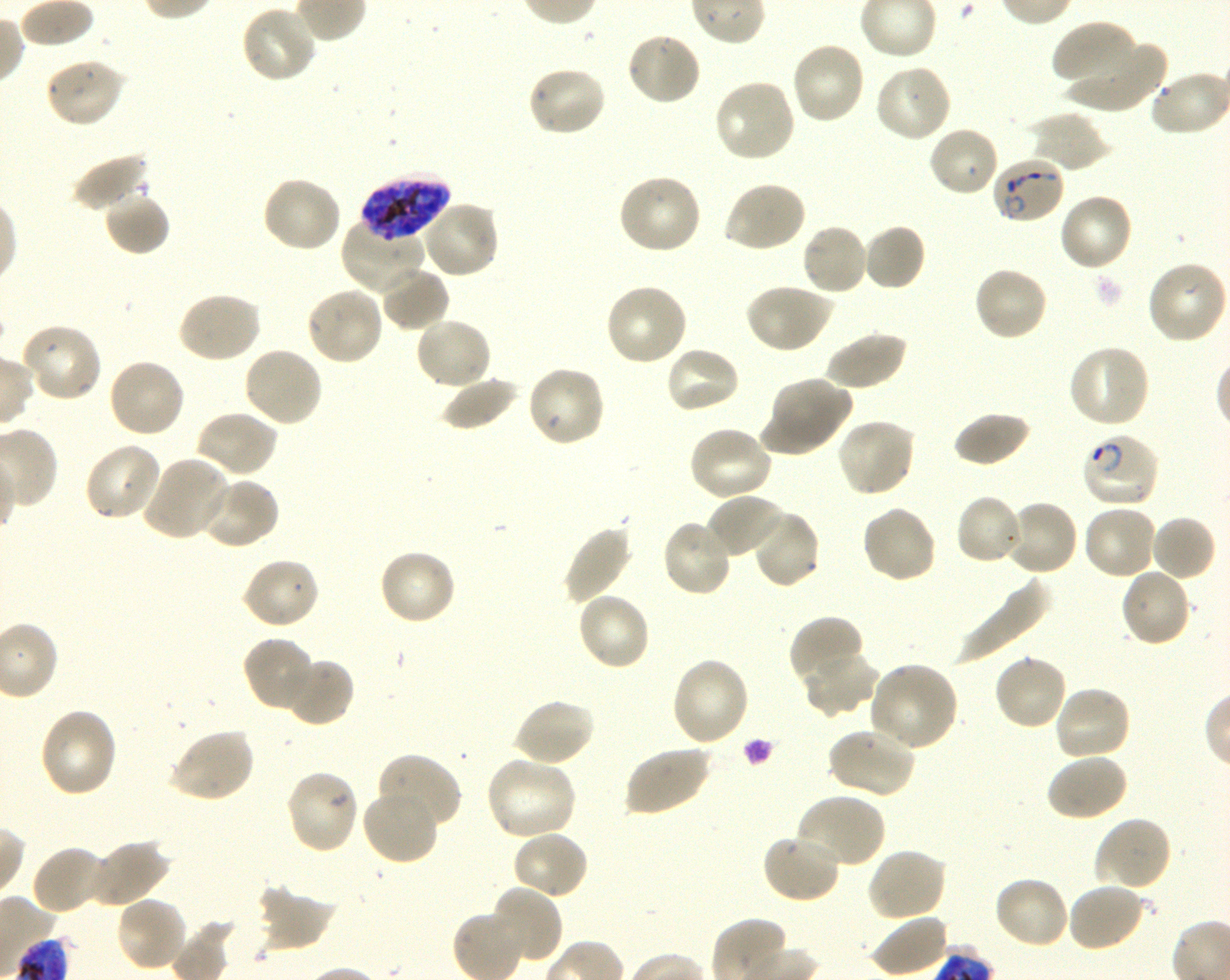

### 4.jpg

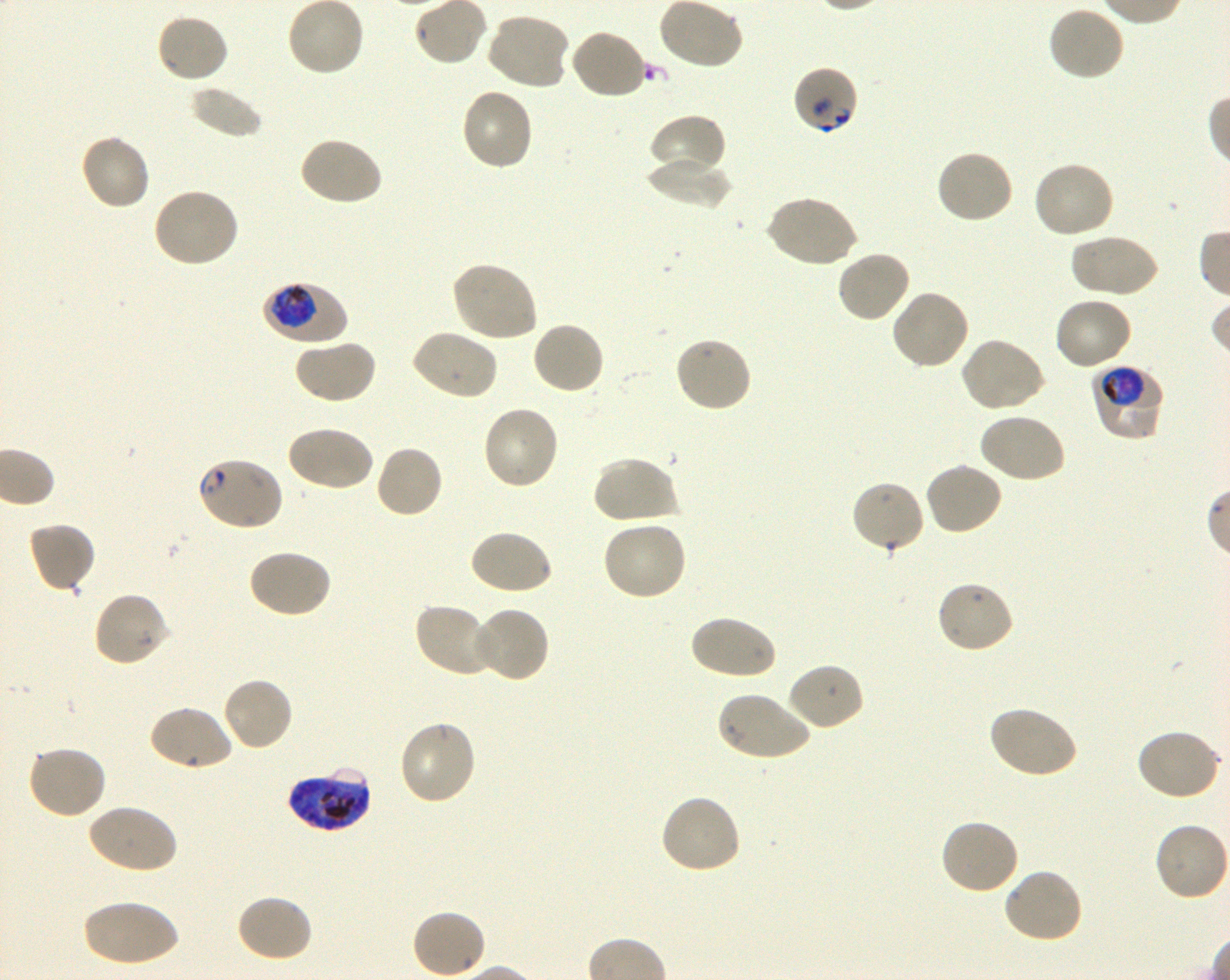

### 5.jpg

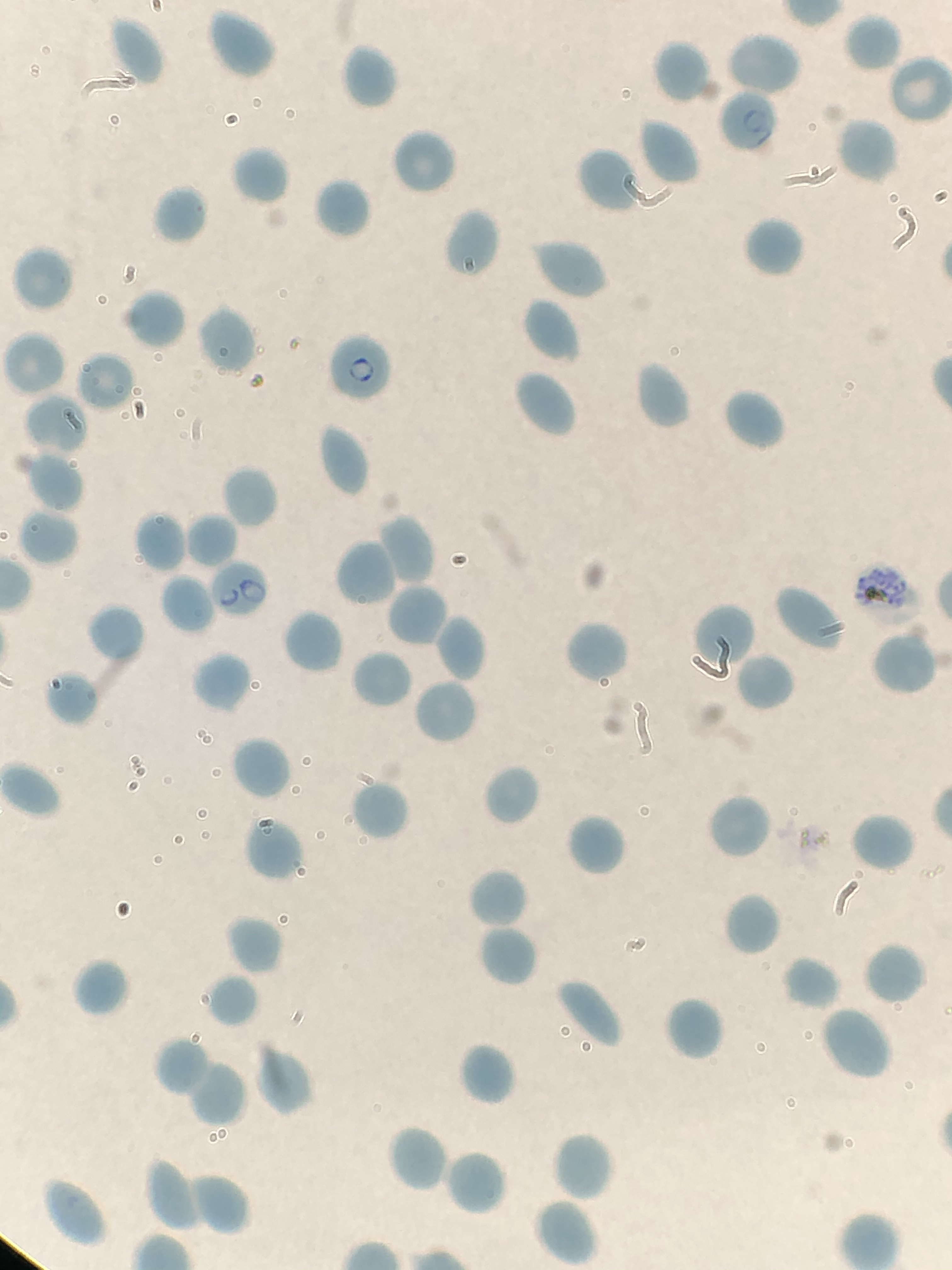

### 6.jpg

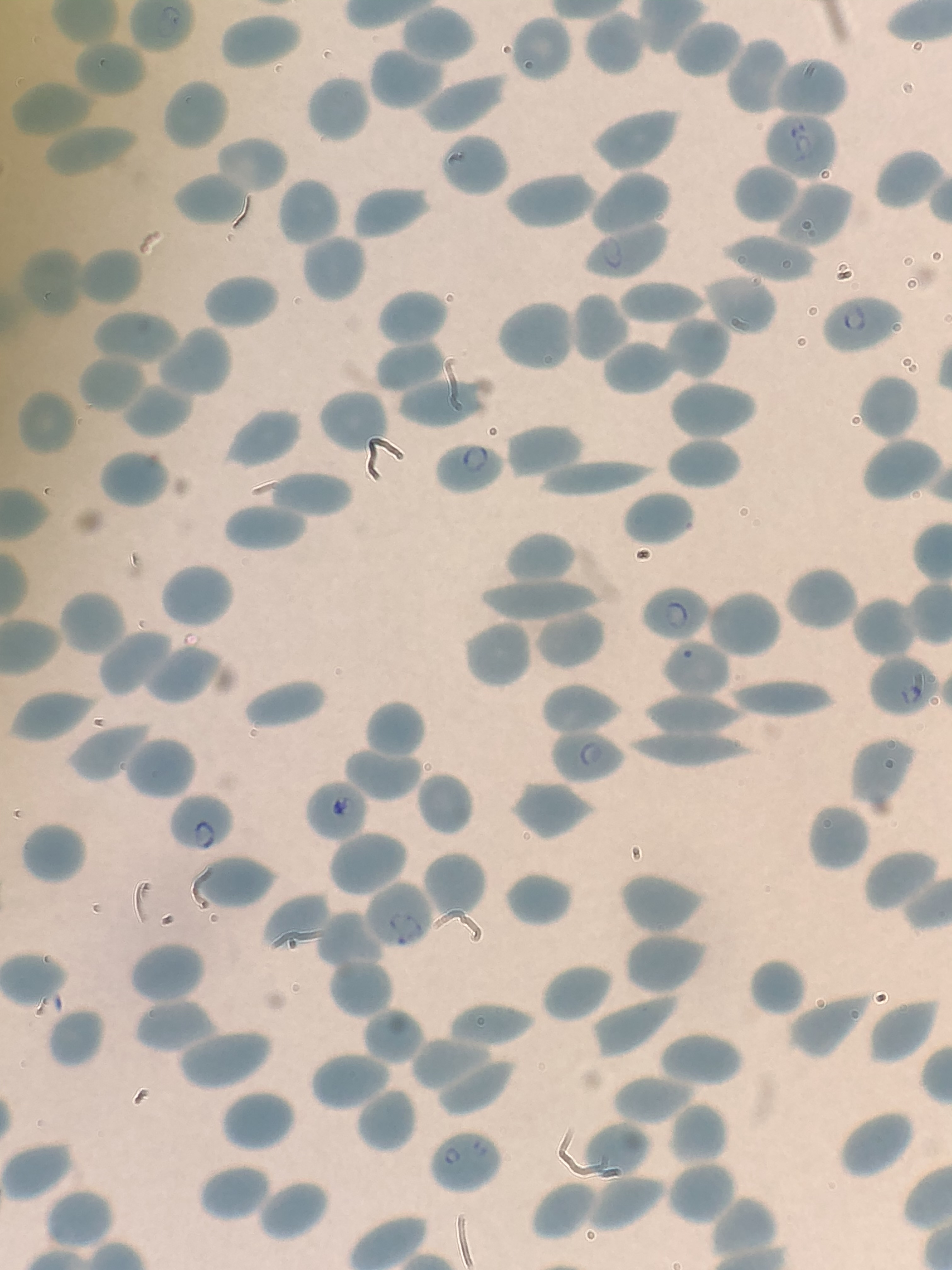

### 7.jpg

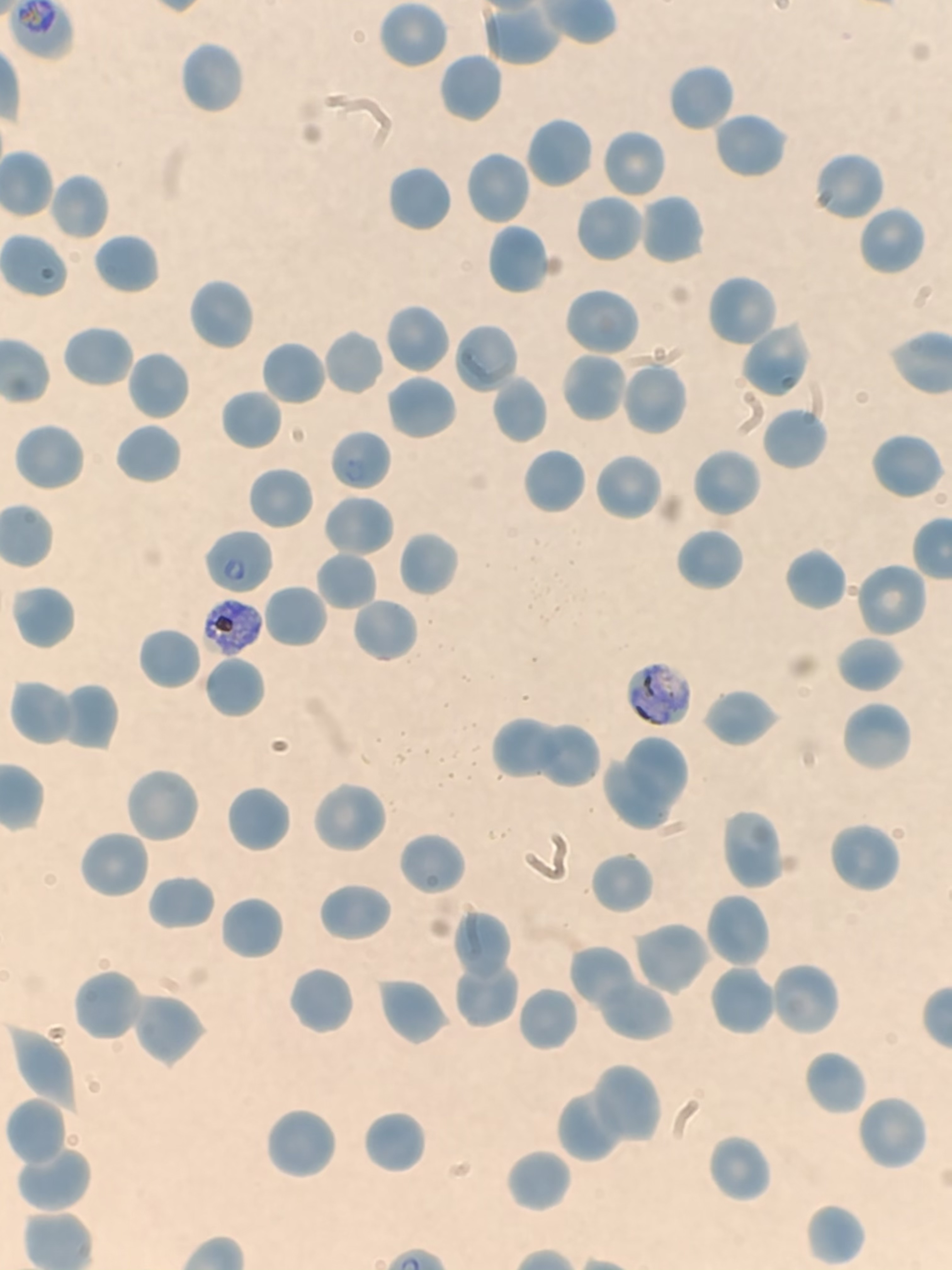

### 8.jpg

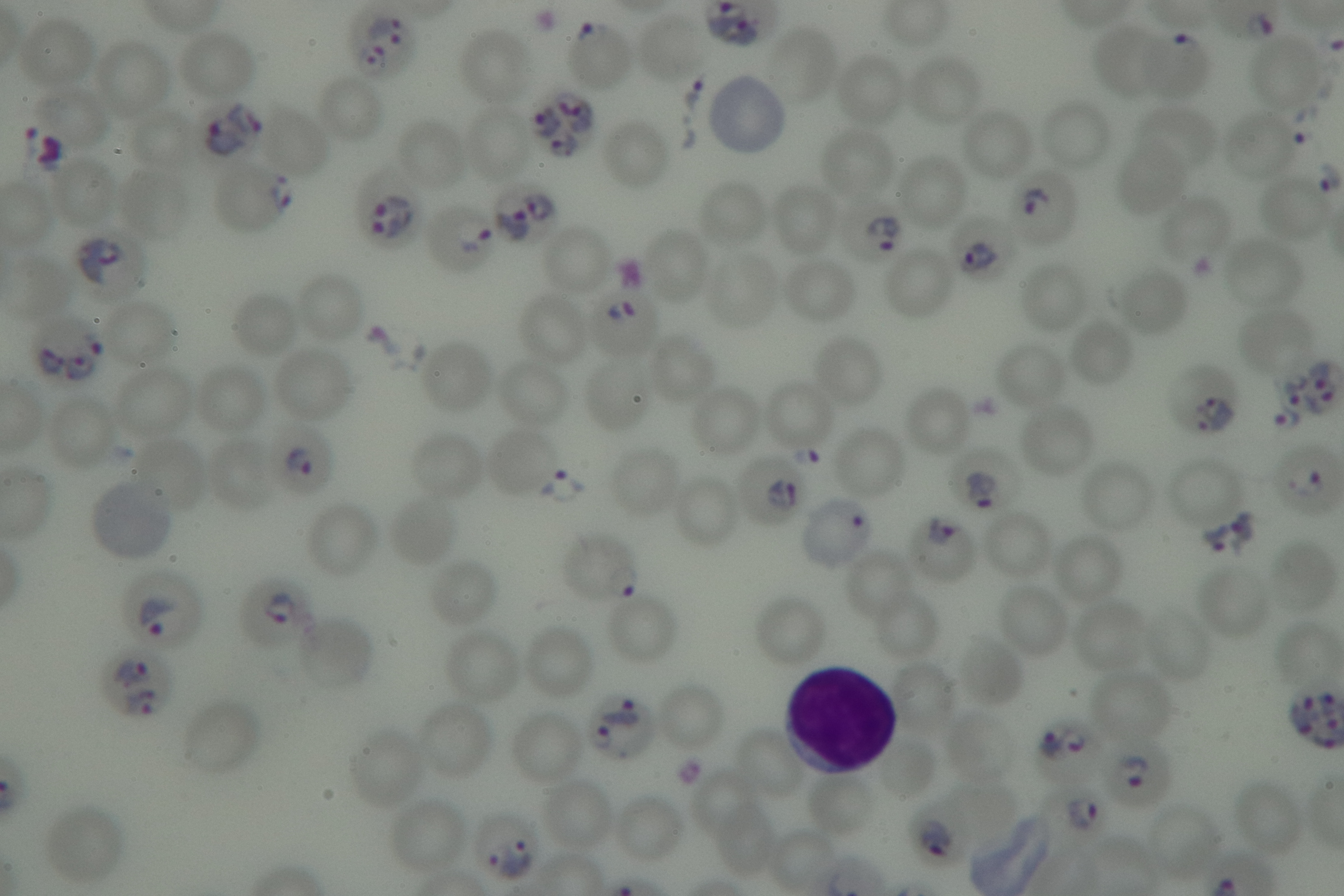

### 9.jpg

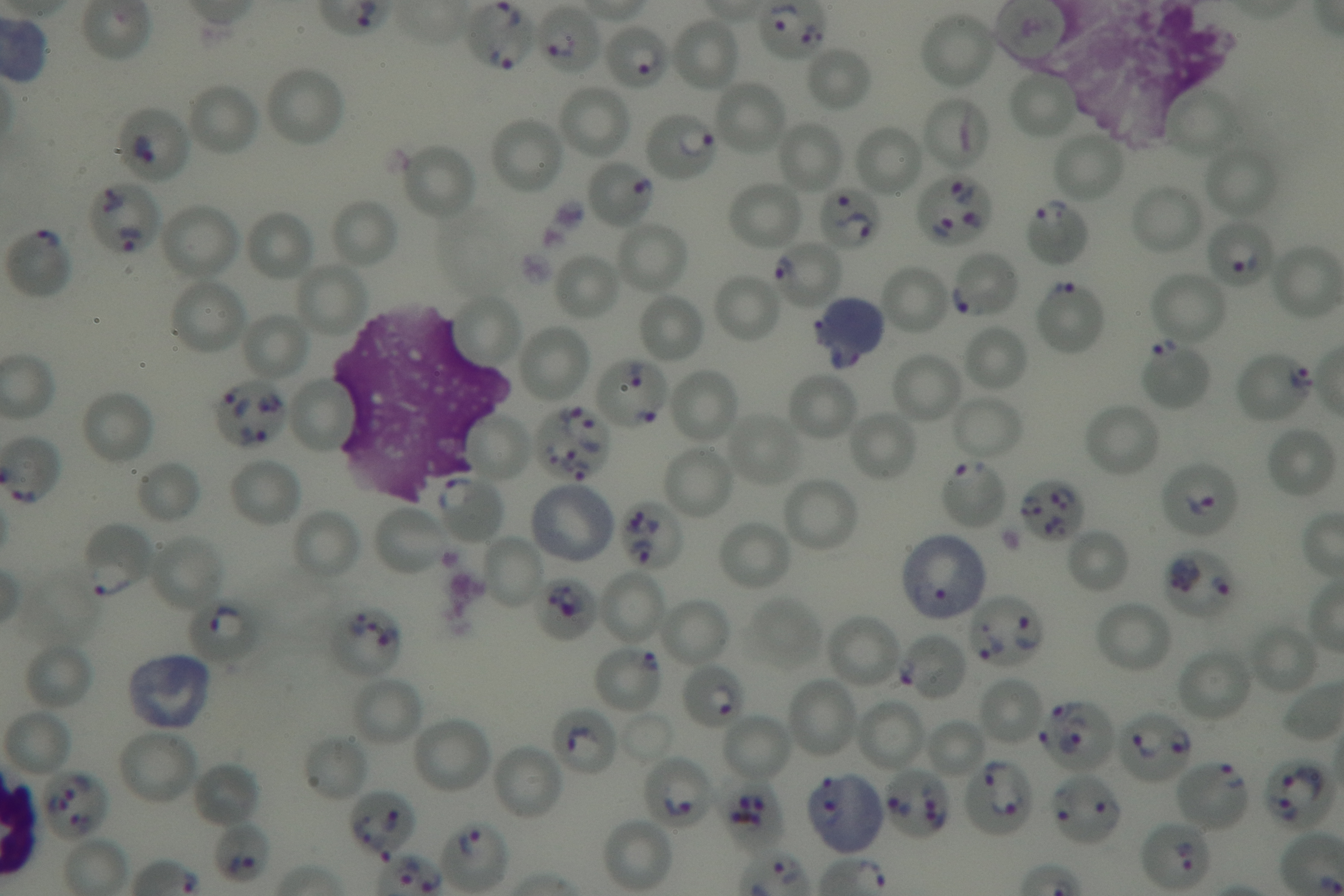
